# Predicting pain and function outcomes in people consulting with shoulder pain: The PANDA-S clinical cohort and qualitative study protocol (ISRCTN 46948079)

**DOI:** 10.1101/2021.05.26.21257208

**Authors:** G Wynne-Jones, H Myers, A Hall, C Littlewood, S Hennings, B Saunders, M Bucknall, S Jowett, R Riley, S Wathall, C Heneghan, J Cook, T Pincus, C Mallen, E Roddy, N.E Foster, D Beard, J Lewis, J Rees, A Higginbottom, DA Van der Windt

**Affiliations:** Primary Care Centre Versus Arthritis, School of Medicine, Keele University; Keele Clinical Trials Unit, School Medicine, Keele University, Staffordshire, UK; Department of Health Professions, Faculty of Health, Psychology & Social Care, Manchester Metropolitan University; Health Economics Unit, Institute of Applied Health Research, University of Birmingham; Nuffield Department of Primary Care Health Sciences, University of Oxford; Department of Psychology, Royal Holloway, University of London; STARS Research and Education Alliance, Surgical Treatment and Rehabilitation Service (STARS), The University of Queensland and Metro North Hospital and Health Service; School of Health and Social Work, University of Hertfordshire, Hatfield AL10 9AB, Hertfordshire, United Kingdom; Department, Central London Community Healthcare National Health Service Trust, London, United Kingdom; Nuffield Department of Orthopaedics, Rheumatology and Musculoskeletal Sciences, University of Oxford

## Abstract

**Introduction:** Shoulder pain is common in primary care but achieving definitive diagnosis is contentious leading to uncertainty in management. To inform optimal primary care for patients with shoulder pain, the study aims to (i) to investigate the short- and long-term outcomes (overall prognosis) of shoulder pain, (ii) estimate costs of care, (iii) develop a prognostic model for predicting individuals’ level and risk of pain and disability at 6 months, (iv) investigate experiences and opinions of patients and healthcare professionals regarding diagnosis, prognosis, and management of shoulder pain.

**Methods and analysis:** PANDA-S is a longitudinal clinical cohort with linked qualitative study. At least 400 people presenting to general practice and physiotherapy services in the UK will be recruited. Participants will complete questionnaires at baseline, 3, 6, 12, 24 and 36 months. Short-term data will be collected weekly between baseline and 12 weeks via SMS text or software application (App). Participants will be offered clinical (physiotherapist) and ultrasound (sonographer) assessments at baseline. Qualitative interviews with ≈15 dyads of patients and their healthcare professional (GP or physiotherapist).

Short and long-term trajectories of shoulder pain and disability (using SPADI) will be described, using latent class growth analysis. Health economic analysis will estimate direct costs of care and indirect costs related to work absence and productivity losses. Multivariable regression analysis will be used to develop a prognostic model predicting future levels of pain and disability at 6-months using penalisation methods to adjust for overfitting. The added predictive value of pre-specified physical examination tests and ultrasound findings will be examined. For the qualitative interviews an inductive, exploratory framework will be adopted using thematic analysis to investigate decision making and perspectives of patients and clinicians on the importance of diagnostic and prognostic information when negotiating treatment and referral options.

**Ethics and dissemination:** The PANDA-S study has ethical approval from Yorkshire and The Humber – Sheffield Research Ethics Committee, UK (18/YH/0346, IRAS Number: 242750). Results will be disseminated through peer-reviewed publications, social and mainstream media, professional conferences, and the patient and public involvement and engagement group supporting this study, and through newsletters, leaflets and posters in participating sites.

**Registration details:** The PANDA-S Study is registered at ISRCTN Number: 46948079

**Article summary:** *Strengths and limitations of this study:*

▪ This cohort study will offer a detailed characterisation of patients presenting with a new episode of shoulder pain in primary care, including short and long-term outcomes.
▪ Detailed, weekly data collection will offer unique insights into the impact of shoulder pain on everyday activity, mood, and work during the first 3 months after presentation.
▪ Clinical assessment will investigate the added predictive value of physical examination tests and ultrasound scan findings, over and beyond self-reported prognostic information.
▪ The use of ‘dyad’ interviews allows for a rich understanding of the views and experiences of clinicians and patients towards shoulder pain management.
▪ The COVID-19 pandemic has impacted on recruitment and data collection, but the study allows an investigation of the pandemic and related (lockdown) measures restrictions on the experience and management of shoulder pain.

## BACKGROUND AND RATIONALE

Shoulder pain is a common condition, with the one-month population prevalence estimated to be between 7% and 26% [1], and an annual incidence in primary care of 29.3 per 1000 person-years.[2] Annually, approximately 3% of adults in the UK will consult their general practitioner (GP) for shoulder pain.[3] The prognosis of shoulder pain is variable with 40-50% of patients reporting persistent pain 6-12 months after first consulting their general practitioner (GP) or physiotherapist [4,5], generating high costs to both healthcare and society.[6-8] Systematic reviews and trials have highlighted modest short-term effects of commonly used treatments such as corticosteroid injection, therapeutic exercise, and manual therapy, but limited evidence for long-term benefits.[9-13]

### Diagnostic uncertainty

Achieving a definitive diagnosis when a patient presents with shoulder pain is contentious and uncertainty exists relating to optimal management based on diagnostic information. Systematic reviews of physical examination tests have noted diversity in performance and interpretation of these tests, poor diagnostic accuracy and lack of evidence about which combination of signs and symptoms are most accurate in predicting patient outcome and response to treatment.[e.g. 14-17] Qualitative research showed that GPs experience uncertainty in the diagnostic work-up of shoulder pain and apply different strategies to deal with uncertainties.[18] There is limited evidence regarding the usefulness of diagnostic imaging, partly due to findings of incidental pathology that does not always correlate with symptom severity.[19,20]

### Prognostic uncertainty

Despite evidence for the prognostic value for a range of factors, it is not clear which combination of prognostic factors optimally discriminates between patients at high versus low risk of poor outcome, with limited evidence for predictive performance of existing prognostic models [21-24] and for their usefulness in routine clinical practice.[25] Short-term symptom change has rarely been investigated but may be highly predictive of long-term outcome, and incorporating monitoring of this early response in the prognostic assessment of individuals with shoulder pain can potentially provide better guidance regarding decisions for further treatment.[26,27] Furthermore, little is known about the pathways that explain favourable or poor outcome in patients with shoulder pain, and generating evidence regarding the role of prognostic factors along these pathways may allow the identification of new targets for treatment.

In summary, given the high impact of shoulder pain, diagnostic uncertainty, variable prognosis, and limited evidence for long-term treatment outcomes, there is a clear need for research investigating short and long-term outcomes of shoulder pain with the aim of improving the primary care management of shoulder pain in future.

#### Aims and objectives

The overall aim of this study is to investigate the short and long-term outcomes of shoulder pain and to develop a prediction model using diagnostic and prognostic information that provides reliable individualised risk prediction for 6-month outcomes of shoulder pain and disability. Specific objectives are to:

1. Describe the short-term (≤6 months) and long-term (up to 3 years) overall prognosis in people presenting with shoulder pain, in terms of pain and function trajectories, and impact on sleep, mood, work, and health-related quality of life.
2. Describe healthcare resource use and estimate the costs associated with care for shoulder pain; and describe and estimate time off work and productivity losses associated with shoulder pain in the short-term (6 months) and long-term (up to 3 years).
3. Develop a prognostic model for predicting individuals’ level and risk of pain and disability at 6 months after presentation, based on self-reported candidate prognostic factors, and estimate and internally validate the model’s predictive performance and clinical utility.
4. Estimate the added prognostic value of physical examination tests and ultrasound scan findings in the prediction of future pain and disability
5. Explore candidate predictors of response to commonly used treatments in a real-life, observational setting
6. Explore perspectives, influences, and uncertainty of patients and clinicians regarding the importance of diagnostic and prognostic information when negotiating treatment and referral options, and making decisions about the management of shoulder pain

## METHODS

### Study design

Multi-centre observational cohort study, including patients presenting with shoulder pain in general practices, and NHS physiotherapy services (including self-referrers to physiotherapy), with a linked qualitative study. Figure details the recruitment methods and participant flow through the study.

#### Patient and Public Involvement

Study questions and design were informed by patient contributors during four dedicated meetings. They highlighted the importance of clear information about the possible cause and prognosis of pain, as this is important to people with shoulder pain when planning their everyday life and considering treatment options. They stressed concern regarding the commonly used approach of ‘watchful waiting’. Postponing treatment/referral decisions was considered frustrating and unhelpful, prolonging the condition and potentially increasing healthcare and personal costs. They expressed the need for a thorough assessment of shoulder pain, along with an early discussion of the possible benefits and drawbacks of diagnostic procedures (e.g. ultrasound scans) and treatment options. The group contributed to the design of the study by advising on recruitment processes, the content of data collection, and how to explain the role of clinical assessment and ultrasound scans to study participants. Annual PPIE meetings are planned to ensure ongoing involvement and engagement during data collection, analysis, interpretation and dissemination of findings.

### Study population

Potential participants will be identified when they consult with an episode of shoulder pain at their general practice, or NHS physiotherapy service (including self-referrers to physiotherapy) in five regions in the UK: Staffordshire, Cheshire, Oxfordshire, Birmingham, and Gloucestershire.

#### Eligibility criteria

Potential participants must be registered at participating general practices, or referred to NHS physiotherapy (including self-referrers), aged 18 years or over, and presenting with a new episode of shoulder pain. A new episode will be defined as no shoulder pain related consultation, no injection, surgery, or physiotherapy-led exercise for shoulder pain, in the last 6 months.

Potential participants will be excluded if they meet the following criteria:

▪ Present to their GP or physiotherapist with symptoms or signs indicative of serious pathology (e.g. fractures, infection)
▪ Have shoulder pain caused by stroke-related subluxation
▪ Have a diagnosis of inflammatory arthritis (e.g. rheumatoid arthritis, or polymyalgia rheumatic)
▪ Have shoulder pain caused by cervical pathology
▪ Are considered by the GP or physiotherapist to be vulnerable (e.g. severe physical and/or mental health problems, dementia).

#### Recruitment

Potential participants consulting with shoulder pain will be identified through one of three methods:

i. Identification using an automated medical record template (pop-up), activated when a Read/SNOMED code for shoulder pain is entered into the medical record as a result of a patient consultation in GP sites.
ii. Identification using an EMIS embedded ‘referral’ form which will auto-populate for eligible patients when triggered by the clinician in physiotherapy sites.
iii. Identification of patients from waiting lists for physiotherapy. Referrals received from GPs, other healthcare professionals, or self-referrals will be centrally triaged by senior physiotherapists using the PANDA-S eligibility checklist.

Potential participants will be asked for consent to share their contact details with the Keele Clinical Trials Unit (CTU) who will send them a study pack, or a study pack will be sent directly from the GP or physiotherapy practices, depending on site preference. The study pack will contain: an invitation letter; a participant information leaflet; baseline questionnaire with consent form and eligibility screening questions; pre-paid reply envelope.

Patients interested to take part in the study will be asked to complete the consent form and answer eligibility questions relating to whether they have received treatment (e.g. supervised exercise, injection, surgery) for their shoulder pain in the 6 months prior to consultation. Those who have not yet received treatment are eligible, and invited to complete and return the baseline questionnaire. As this is an observational study, all participants will continue to receive care as usual for their shoulder pain.

### Data collection

Data collection will be carried out by postal questionnaire at baseline, 3, 6, 12, 24, and 36 months. Since June 2020, in response to the COVID-19 pandemic, follow-up questionnaires have also been made available as online surveys. Short term data will be collected weekly through a specifically designed smartphone/tablet Shoulder Pain App or text messages over 12 weeks. Clinical data will be collected during a clinical assessment comprising a shoulder examination by a physiotherapist and an ultrasound scan. In response to the pandemic, face to face clinical assessments are carried out (or paused) in accordance with national and local restrictions. Table 1 provides an overview of the content and timing of data collection for this study.

**Table 1.** Data collection schedule (self-report questionnaires)

| Description | Measure (no. of items, response options, score range) | Baseline<br>(shortly after consultation) | 3 months | 6 months | 12 months | 24 months | 36 months |
| --- | --- | --- | --- | --- | --- | --- | --- |
| Primary outcome measure |  |  |  |  |  |  |  |
| Severity of pain | SPADI total score (5 pain, 8 disability items, 0-10 NRS, 0-100) | ✓ | ✓ | ✓ | ✓ | ✓ | ✓ |
| Severity of disability |  | ✓ | ✓ | ✓ | ✓ | ✓ | ✓ |
| Secondary outcome measures |  |  |  |  |  |  |  |
| Sleep | Jenkins sleep questionnaire (4 items, 3 options each, 0-8) | ✓ | ✓ | ✓ | ✓ | ✓ | ✓ |
| Work absence | How many days off work have you had in the past month (days/weeks) | ✓ | ✓ | ✓ | ✓ | ✓ | ✓ |
| Work performance | Single item question (0-10) | ✓ | ✓ | ✓ | ✓ | ✓ | ✓ |
| Global perceived change | Single item question (6 options) | ✗ | ✓ | ✓ | ✓ | ✓ | ✓ |
| Health-related quality of life | EQ-5D-5L (5 items, 5 options each) | ✓ | ✓ | ✓ | ✓ | ✓ | ✓ |
| Socio-demographics |  |  |  |  |  |  |  |
| Age | Date of birth | ✓ | ✗ | ✗ | ✗ | ✗ | ✗ |
| Gender | Male / Female / Prefer not to say | ✓ | ✗ | ✗ | ✗ | ✗ | ✗ |
| Education | Highest qualification (single item, 5 options) | ✓ | ✗ | ✗ | ✗ | ✗ | ✗ |
| Health literacy | Single item question (5 options) | ✓ | ✗ | ✗ | ✗ | ✗ | ✗ |
| Work status | 3 items | ✓ | ✗ | ✗ | ✓ | ✓ | ✓ |
| Shoulder pain characteristics |  |  |  |  |  |  |  |
| Side involved | Left / right / both | ✓ | ✗ | ✓ | ✓ | ✓ | ✓ |
| History | Number of episodes in both shoulders (2 items, 4 options) | ✓ | ✗ | ✗ | ✗ | ✗ | ✗ |
| Onset | Acute versus gradual (2 options) | ✓ | ✗ | ✗ | ✗ | ✗ | ✗ |
| Duration | Single item question (6 options) | ✓ | ✗ | ✗ | ✗ | ✗ | ✗ |
| Which is your dominant arm | Left / Right | ✓ | ✗ | ✗ | ✓ | ✓ | ✓ |
| Continued shoulder pain at follow-up | Two single item questions | ✗ | ✗ | ✗ | ✓ | ✓ | ✓ |
| Pain elsewhere | Full body manikin | ✓ | ✗ | ✗ | ✗ | ✗ | ✗ |
| Previous imaging for shoulder pain | Pre-defined list (4 options) |  |  |  |  |  |  |
| Previous shoulder pain treatments | Pre-defined list (6 options) | ✓ | ✗ | ✗ | ✗ | ✗ | ✗ |
| Analgesic use (over the counter and GP prescribed) | predefined list (10 options) | ✓ | ✓ | ✓ | ✓ | ✓ | ✓ |
| **Comorbidities and lifestyle** |  |  |  |  |  |  |  |
| Comorbidity | Diabetes, insulin-dependent or not (3 options) | ✓ | ✗ | ✗ | ✓ | ✓ | ✓ |
| Height and weight (BMI) | Self-reported | ✓ | ✗ | ✗ | ✓ | ✓ | ✓ |
| Smoking and vaping (frequency) | Two single item questions (4 options each) | ✓ | ✗ | ✗ | ✓ | ✓ | ✓ |
| Alcohol consumption (frequency) | Single item question (6 options) | ✓ | ✗ | ✗ | ✓ | ✓ | ✓ |
| Physical activity (frequency) | Single item question (7 options) | ✓ | ✗ | ✗ | ✓ | ✓ | ✓ |
| **Work related factors** |  |  |  |  |  |  |  |
| Most recent paid job title | Single item question | ✓ | ✗ | ✗ | ✓ | ✓ | ✓ |
| Current work situation | Single item question | ✓ | ✗ | ✓ | ✓ | ✓ | ✓ |
| Psychosocial work environment | Single component of the Work Organisation Assessment Questionnaire (4 items, 5 options, 0-16) | ✓ | ✗ | ✓ | ✓ | ✓ | ✓ |
| Attitudes and beliefs towards work | Newly developed questionnaire, 11 items, 7 options, 0-66) | ✓ | ✗ | ✓ | ✓ | ✓ | ✓ |
| **Psychosocial and behavioural factors** |  |  |  |  |  |  |  |
| Anxiety and depression | Hospital Anxiety and Depression Scale (14 items, 4 options each, 0-42) | ✓ | ✓ | ✓ | ✓ | ✓ | ✓ |
| Fear of moving the arm | Single item question (0-10 NRS) | ✓ | ✓ | ✓ | ✓ | ✓ | ✓ |
| Pain self-efficacy | Pain Self Efficacy Questionnaire (10 items, 7 options each, 0-60) | ✓ | ✓ | ✓ | ✓ | ✓ | ✓ |
| Consultation-based reassurance | Consultation-based reassurance questionnaire, 12 items, 7 options, 4 subscales 0-28) | ✓ | ✓ | ✓ | ✗ | ✗ | ✗ |
| Worry about shoulder pain | Single item question (0-10 NRS) | ✓ | ✓ | ✓ | ✓ | ✓ | ✓ |
| Treatment expectations (confidence) | List of potential treatment options (7 items, 0-10 NRS) | ✓ | ✗ | ✓ | ✓ | ✓ | ✓ |
| **Health economic measures** |  |  |  |  |  |  |  |
| Healthcare utilisation for shoulder pain (prescriptions, GP consultations, investigations referrals) | Self-reported (follow-up questionnaires; 4 items) and medical record review | ✓ | ✗ | ✓ | ✓ | ✓ | ✓ |

NRS: numerical rating scale

#### Primary outcome measure

The primary outcome measure for investigating course, prognosis, and treatment response is the Shoulder Pain and Disability Questionnaire (SPADI).[28] The SPADI is scored using a 0-10 numerical rating scale for each question from “no pain” to “worst imaginable pain” (for the pain scale) and from “no difficulty” to “so difficult that help is required” (for the disability scale). Numerical scores are summated and divided by the maximum score possible for all relevant questions and then multiplied by 100 to generate a score from 0-100 with higher scores indicating worse shoulder pain and/or disability.

#### Secondary outcomes

Pain intensity will be measured on a numerical rating scale from 0-10 asking the participant to report their worst pain in the past week. Sleep will be measured using the Jenkins Sleep Questionnaire.[29] Global perceived change in shoulder pain since the baseline questionnaire will be asked in all follow-up questionnaires with one question providing participants with six possible response options from “completely recovered” to “much worse”. Work absence will be assessed through two methods:

▪ Employed participants will be asked if they are currently absent from work due to their shoulder pain, and, if so, for how long they have been absent
▪ Fit note data for shoulder pain will be collected from the medical record (where participants have provided consent) allowing data on the number of days and number of episodes of clinician certified work absence to be collected.

Participants will also be asked to indicate how their shoulder pain has impacted on their work performance using the question “On average, to what extent has pain affected your performance at work in the past x months” [30] and are asked about attitudes and beliefs regarding health and work, measured using a new developed set of 11 items.

Healthcare utilisation will be estimated based on self-report (follow-up) questionnaires, and include primary care consultations (general practitioners and practice nurses), secondary care consultations (e.g. hospital consultants, physiotherapists), prescriptions, hospital based procedures (diagnostic tests, injections, and investigations) nature and length of inpatient stays, and surgery. Patients will be asked to distinguish between UK NHS and private provision. Finally, the EQ-5D-5L [31] will be used to measure participants’ health-related quality of life.[32]

#### Candidate prognostic factors and moderators

Questionnaires will measure self-report candidate prognostic factors and predictors of treatment outcome (objectives 1,3 and/or 5) which have been selected based on previous systematic reviews and cohort studies:

▪ Sociodemographic variables: age, gender, level of education, health literacy [33]
▪ Shoulder pain characteristics: history, duration, onset, and baseline severity of shoulder pain/disability; pain elsewhere (full body manikin)[34]
▪ Co-morbidities and lifestyle: diabetes and other relevant long-term conditions, height and weight to calculate body mass index (BMI), smoking and vaping, alcohol consumption, and physical activity
▪ Work-related factors: current work status
▪ Psychosocial and behavioural factors:
  ⍰ Symptoms of anxiety and depression (Hospital Anxiety and Depression Scale (HADS) [35]
  ⍰ Fear-avoidance beliefs, derived from the Fear-Avoidance and Beliefs Questionnaire [36], and worry about shoulder pain measured using single item questions
  ⍰ Pain self-efficacy (Pain Self Efficacy Questionnaire (PESQ)[37])
  ⍰ Cognitive and affective reassurance (Consultation-based Reassurance Questionnaire [38,39]
  ⍰ Treatment expectations: questions asking participants how confident they are that specified treatments will help their shoulder pain.

During the COVID-19 pandemic it is anticipated that participants’ responses to some questions may be influenced by the pandemic or related restrictions, particularly questions relating to current work status, ability to participate in usual activities, shoulder pain treatments, self-management of shoulder pain, anxiety, and depression. To capture this, participants will be given the opportunity to comment on any conditions or circumstances that may have affected their responses in the follow-up questionnaires (open-ended question).

#### Medical record review

Participants will also be asked for consent to access and export aspects of their medical records, to provide information on healthcare resource use including information on fit notes; prescriptions; consultation frequency; referrals for further treatment and procedures (e.g. imaging, surgery); non-shoulder-related musculoskeletal consultations and other relevant comorbidity (coronary heart disease, diabetes, thyroid disease, cancer).

#### Follow-up data collection

Participants will be sent follow-up questionnaires at 3, 6, 12, 24 and 36 months after the return of their baseline questionnaire; these follow-up questionnaires will be sent by post or a link to an online questionnaire will be sent by email. Non-responders to follow-up questionnaires will receive a reminder questionnaire after 2 weeks, 4 weeks, and a telephone call a further 2 weeks later if no response has been received, with the option of completing a short (Minimum Data Collection (MDC)) questionnaire by telephone, and if no telephone contact can be made, by post.

#### Short term data collection

On return of the baseline questionnaire participants will be offered the option of completing the shoulder pain app or text messages reporting their pain and function (0-10 Numerical Rating Scale, NRS) weekly for 12 weeks. The app also collects weekly data across 8 further domains using single item questions: self-efficacy, work absence, mood, sleep, medication use, fear of movement, worry, treatment, and recovery expectations.

### Clinical assessment

Those participants who have returned a completed questionnaire will be offered the option to attend a clinical assessment by an experienced and trained physiotherapist. Participants will be notified that all information collected at the clinic will be collected for research purposes, and that no recommendations will be given regarding diagnosis, treatment, or referral. Findings from the clinical assessment will only be discussed with their GP if the assessor feels there is a need for immediate clinical attention. In response to restrictions imposed during the coronavirus pandemic, ethical approval was requested (as an amendment to the original approval) to share a brief report from the ultrasound scan with the participants’ GPs. This may avoid the need for a separate referral for ultrasonography for clinical purposes in a context where there is a need to reduce physical contact and travel for non-essential purposes.

#### Physical examination

The assessment will include a standardised history based on questions regarding duration, severity, impact, and (self) management of shoulder pain. The physical examination will include an examination of the neck using repeated movements through flexion, extension, and side-flexion, in order to assess whether the patient’s shoulder pain is related to a neck problem. Shoulder range of movement will be assessed during active abduction, internal and external rotation, compared to the contralateral side. Additional tests designed to distinguish between different shoulder conditions have variable sensitivities and specificities, but was informed by recent systematic reviews [1416,17]:

▪ Painful arc, Neer sign and Hawkins-Kennedy tests
▪ External rotation lag sign
▪ Glenohumeral external rotation
▪ Scapular assistance test
▪ Empty Can and Full Can test
▪ Scarf test and Bear Hug test
▪ Step-standing elevation
▪ Muscle performance tests aiming to identify weakness and pain

Anthropometric measurements will include height (in centimetres - cm) and weight (in kilograms - kg) and waist-hip circumference (in cm). To assess balance, strength, and mobility, we will include a brief standardised protocol, consisting of grip strength, lower limb strength (sit-to-stand) and a balance test.

Following the assessment, the research physiotherapist will record their opinion regarding the pathoanatomical classification of the shoulder problem (rotator cuff disorder, frozen shoulder, glenohumeral osteoarthritis, acromioclavicular joint pain, instability, neck-related dysfunction), and rate their confidence in this classification using a 0-10 NRS. The physiotherapist will also estimate the participant’s prognosis, through answering the question “Do you think this participant will have interfering shoulder pain in 6 months’ time? Yes/No/Don’t know”.

#### Ultrasound assessment

The ultrasound scans (US) will be performed by experienced ultrasonographers/radiologists using high resolution ultrasound systems and transducers [40] according to a standardised scanning protocol.[41] The structures scanned will include long head of biceps tendon, rotator cuff (subscapularis, supraspinatus, infraspinatus and teres minor tendons and muscles), posterior glenohumeral joint, subacromial/deltoid bursa, and acromioclavicular joint. A standardised structured report based on [42] will include information regarding tendon and bursal pathology including structural appearances, site, and size of tendon tears, observed glenohumeral or acromioclavicular fluid, synovitis or cortical bone changes and muscle atrophy. Dynamic scanning of some shoulder movements will be performed to assess for restriction or instability. Colour Doppler will be used to assess the presence of neovascularisation in any areas of tendon, sheath or bursal abnormality. Both shoulders will be scanned to enable later analysis of abnormalities in affected versus pain-free shoulders.

### Qualitative interview study

#### Recruitment of patient-clinician dyads

Patient-participants: Questionnaires will be screened to enable a purposive sampling frame to be applied. A range of participant characteristics will be sampled for, including age, sex, reported pain intensity, pain duration, shoulder diagnoses, socioeconomic status, health literacy, and Fit Note status (i.e. absent from work or not). Participants who are selected for interview will be sent an invitation letter, reply slip and participant information leaflet about the interview. Those who return a reply slip indicating a willingness to be interviewed will be telephoned to arrange the interview.

Clinician-participants: As part of the patient-participant consent process, the participant will be asked the name of the clinician with whom they consulted for their shoulder pain (GP or physiotherapist) and for permission to contact the clinician to arrange a separate individual interview in which the consultation will be discussed. If a participant declines consent to contact their clinician, then the participant’s interview will be used alone.

#### Data collection

Topic guides will be used in interviews, which will be informed by the study objectives and the future aim of designing an optimal (stratified) model of care for shoulder pain, and by previous research on participant-clinician communication, effective reassurance, and diagnostic and prognostic uncertainty in musculoskeletal pain.[38,43,44] Separate topic guides are developed for participant and clinician interviews. Clinicians will be asked about their views and experiences of treating shoulder pain, particularly in relation to the given consultation. Topic guides will be iteratively revised throughout the data-collection process in light of emergent findings.

Topics will include (but not be limited to):

▪ Participants’ experiences of managing their shoulder pain condition, and its impact upon their lives
▪ Participants’ understanding of possible causes of shoulder pain, including the ‘label’ attached to explanations and identified issues associated with this
▪ The value participants and clinicians attribute to diagnostic tests, including physical examination and imaging
▪ Consideration of explanations, concerns, and uncertainty regarding prognosis
▪ Views on decisions and advice given about self-management, work and other activities.
▪ Clinician-patient communication regarding diagnosis, prognosis and treatment options.
▪ Participants’ and clinicians’ views and experiences regarding the impact of the coronavirus pandemic on management of shoulder pain and on treatment and referral decisions.

Semi-structured interviews will be carried out with approximately 15 participant-clinician dyads (i.e. approximately 30 interviews in total). The final number of interviews will be guided by data saturation, defined in terms of ‘informational redundancy’ [45] [the point at which additional data no longer offers new insights.

### Analysis cohort study (objectives 1-5)

A detailed analysis plan will be written for each of the study objectives, a brief summary is given here.

#### Objective 1: Investigate pain and function trajectories (overall prognosis)

Descriptive statistics will be used to report baseline characteristics of the study population, and the course of symptoms over time, for the primary outcome (SPADI) and secondary outcome measures. For each follow-up time-point (0,3,6,12,24,36 months) we will report means for continuous outcomes, and proportions for binary outcomes, based on longitudinal models that account for correlated responses over time. Attrition (n,%) will be described for each follow-up time-point. Baseline characteristics among those lost during follow-up (whose outcomes are hence not fully observed) will be summarized and compared to characteristics of those remaining in the study to assess for risk of attrition bias.

Latent class growth analysis or other latent trajectory analysis will be used to identify distinct groups of participants with similar short-term trajectories of shoulder pain and function scores (0-10 NRS) using weekly measurements of outcome data at up to 12 time-points over the first 3 months. For inclusion in the analysis, participants will be required to have available data in week 1 and at ≥2 further time-points. The optimal number of trajectories will be selected using a combination of statistical, parsimony and interpretability criteria.[46]

Appropriate polynomial functional form for each trajectory will be chosen, based on the significance of the estimated parameters related to each polynomial component and participants will be assigned to trajectories according to maximum probability assignment principle. Number of trajectories will be increased and model fit assessed using Bayesian Information Criterion (BIC) and sample size adjusted BIC. The Lo, Mendell and Rubin adjusted likelihood ratio test (LRT) [47], and the bootstrap LRT were used to assess whether there was a significant improvement in model fit between *k-1* and *k* trajectory models. The following criteria will also be used for model selection: (a) delineation of trajectories assessed by higher entropy, (b) average posterior probability of trajectory membership >0.7, (c) trajectory membership ≥4% and (d) clinical relevance and interpretation of the identified trajectories.

Baseline characteristics of subgroups showing distinct trajectories will be described, including any treatment received for shoulder pain during the first three months. Similar methods will be used to describe long-term trajectories, using outcome data at 6, 12, 24, and 36 months. Based on previous studies investigating trajectories in other conditions (n=350-700) [e.g. 48,49] we expect to identify between 3 and 5 classes, for which a sample size between 400 and 500 participants should be sufficient. The GRoLTS checklist for reporting latent trajectory studies will be followed.[50]

#### Objective 2: Describe healthcare resource use and productivity losses

Health care costs will be estimated by combining resource use data with unit costs, obtained from standard sources including the British National Formulary (BNF) for drugs[51], NHS Reference costs [52] and Unit Costs of Health and Social Care.[53] Productivity costs will be estimated using the human capital approach with salary costs based on respondent job-specific average wage estimates identified from annual earnings data and UK Standard Occupational Classification coding.[54] Responses to the EQ-5D-5L at each time point will be converted to utility values obtained using the crosswalk value set, in line with current National Institute for Health and Care Excellence (NICE) recommendations.[55] Utility values will also be converted into quality-adjusted life years (QALYs) using the area-under-the-curve approach linking utility scores at various time-points. A descriptive analysis of resource use, health care costs, time off work, productivity costs, EQ-5D-5L utility values and total QALYs will be conducted, with presentation of means and confidence intervals obtained by non-parametric bootstrapping.

#### Objective 3: Develop a prognostic model for predicting individuals’ level and risk of pain and disability

We will develop and validate a multivariable prediction model for reliably estimating expected levels of pain and disability (using SPADI total score as a continuous outcome) over 6 months follow-up. Development of prognostic prediction model will be guided by the PROGRESS framework [56-58], and reported using TRIPOD reporting guidelines. The number of variables (candidate predictors) for potential inclusion will be restricted to meet sample size requirements (see below), based on existing or emerging evidence regarding the prognostic value of variables, combined with clinical and patient expertise, and consider sociodemographic variables, lifestyle factors, shoulder pain characteristics, comorbidity, and psychosocial factors. Candidate predictors will be based on participant self-report to allow wide application of the prognostic model, with data extracted from the baseline questionnaire. Multivariable linear regression using the elastic net penalty (to penalise for potential overfitting and to allow variable selection) Continuous prognostic variables will not be categorised during the process of developing the models, with splines or fractional polynomials used to examine non-linear associations with outcome. Prognostic subgroups will be defined based on predicted SPADI values at 6 months, using a priori defined thresholds for SPADI for recovery (eg. SPADI score <20) or persistent high levels of pain and disability (e.g. SPADI score ≥50). We will additionally describe predictive performance of the model to accurately predict the probability of individuals with shoulder pain to experience recovery and risk persistent high SPADI scores. Internal validation will be undertaken using bootstrapping of the entire development dataset, and optimism-adjusted estimates of predictive performance produced for calibration (e.g. R^2^, calibration-in-the-large, calibration slope) and discrimination (e.g. C-statistic, area under the curve) for predicted risks.

We have estimated minimum sample size using the approach proposed by Riley et al. [59,60], and using the Stata pmsampsize command, aiming to reduce overfitting of the prediction model. Based on previous studies, we expect R-squared to be 0.5 when including baseline level of the primary outcome (SPADI total score) in the model as planned. Based on an expected mean change of 25 points and SD of 23 [5] and a model including 20 candidate predictors, the required minimum sample size is 254 participants. Accounting for loss to follow-up of 25% we need to recruit at least 339 participants to the cohort. When using a binary outcome, a minimum sample size of 377 participants with follow-up (503 at baseline) would be needed.

#### Objective 4: Explore the added prognostic value of physical examination tests and ultrasound scan findings

Due to lockdown measures during the COVID-19 pandemic, a much smaller number of participants than originally planned can be invited for a clinical assessment. Given the expected small sample size, the statistical analysis of data from the clinical assessment will focus on testing a priori defined hypotheses, informed by previous literature and clinical expertise, and explore the value of adding specific physical examination tests or ultrasound scan findings for predicting outcome (higher SPADI scores over 6 months follow-up), over and above prognostic information included in the prediction model using self-report data only. Assuming R-squared from the prediction model will be 0.5 using 20 predictors (as described above) in order to have 80% power to detect one additional predictor adding 0.05 to the R-squared to the model, we would need 74 participants without drop-out, and 99 participants assuming 25% will drop-out.[61]

#### Objective 5: Explore candidate predictors of response to commonly used treatments in a real-life, observational setting

Developmental work (systematic review, workshops with clinicians, and international choice-based conjoint-survey of clinicians)[62] has generated a shortlist of candidate treatment moderators that may modify the response to commonly used treatments for shoulder pain in primary care (advice and pain relief only, corticosteroid injection, exercise/mobilisation), as recorded by participants in the three and six months follow-up questionnaires. These candidate moderators include symptom duration; presumed cause of shoulder pain (injury or other); co-existing neck pain; psychosocial complexity (fear-avoidance, catastrophizing, anxiety, depression); positive expectations or preferences regarding treatment; comorbidity (in particular diabetes). Linear (random effects) regression analysis will be used to estimate the outcomes of treatment received (e.g. corticosteroid injection compared to advice/analgesics only), using SPADI score over 6 months follow-up as the primary outcome. Propensity score methodology will be used to adjust for confounding by indication due to observed covariates. Sensitivity analysis will be performed to assess the robustness of findings to potential unmeasured confounding, using E value approach.[63] The effects of moderator*treatment interactions will be explored, and overall treatment outcomes as well as outcomes for relevant subgroups (where relevant moderator*treatment interactions will be identified) described. Based on previous primary care studies [7] and our GP survey [64] we expect that 20-40% of patients receive an injection and 25-50% of patients see a physiotherapist following GP consultation. Depending on the distribution of candidate predictors, this will give subgroups of ≥100 to explore the role of candidate moderators. These are exploratory analyses, given that predictors of treatment effect need to be confirmed using data from randomized trials, but will offer insight into their value in the broader population of people with shoulder pain presented in routine primary care.

#### Missing data

For all quantitative analyses patterns of missing data will be described. Under a ‘missing-at-random’ assumption, individuals with partially missing outcome data (e.g. at some time-points) will be included in analyses (without imputation) using a longitudinal data (hierarchical) modelling framework. If there is a considerable amount of missing baseline data for candidate predictors or covariates of interest this will be handled using multiple imputation, and Rubin’s Rules used to combine results across imputed datasets. The imputation will be conditional on observed outcomes and candidate predictors, and also auxiliary variables, to help ensure a ‘missing-at-random’ assumption is appropriate.

### Analysis qualitative study (objective 6)

All interviews will be audio-recorded, fully transcribed and then cleaned and anonymised. An inductive, exploratory framework will be adopted using thematic analysis, and influenced by grounded theory.[65] The constant comparison method [66] will be used in the analysis, looking for connections within and across interviews, and across codes, highlighting data consistencies and variation.

The participant-clinician dyad will be the unit of analysis. Exploring dual perspectives on the consultation can provide a rich data source which can strengthen trustworthiness of data [67]. Comparisons will be made between the matched participant and clinician interviews, looking for similarities and differences in the separate accounts given. Analysis will therefore draw comparisons both between clinician and participant perceptions of specific consultations as well as between different clinician-participant dyads across the sample as a whole.

## DISCUSSION

This study protocol describes a prospective cohort study investigating the course and prognosis of shoulder pain in primary care, as well as healthcare costs and productivity losses associated with an episode of shoulder pain. The study includes a linked qualitative study, interviewing dyads of patients and their clinicians about the influences on decision making and their perspectives on the importance of diagnostic and prognostic information in the management of the shoulder problem. Recruited is expected to be completed in July 2021.

The cohort study is part of a programme of work aiming to improve patient outcomes and healthcare resource use by early, more effective targeting of patients to treatments from which they are likely to benefit most. The cohort will provide insight into the content and outcomes of current primary care for patients presenting with shoulder pain, describe overall prognosis, and use self-report information from participants to develop a prognostic model for predicting future levels of pain and disability in individuals with shoulder pain, allowing early identification of those who can be reassured and self-manage their shoulder condition, as well as those who are at risk of persistent pain and disability and would benefit from further treatment. The PANDA-S research programme also includes an Individual Participant Data (IPD) meta-analysis of randomised trials to test candidate predictors of the effect of commonly used treatments for shoulder pain.[68] The data from the cohort study, linked qualitative study, and IPD meta-analysis will then be used to co-design, together with clinicians and patients, a prognostic screening and treatment decision tool to inform improved decision-making for people presenting with shoulder pain. The use of the tool will subsequently be evaluated in a multi-centre, pragmatic randomised trial.

The coronavirus pandemic, including restrictions required to reduce the risk of infection, has impacted significantly on the plans for data collection in the cohort study, limiting the number of patients that can be offered a clinical assessment and ultrasound scan within the timeframe of the study. The statistical analysis of physical examination tests and ultrasound scan findings will therefore focus on testing specific hypotheses regarding the prognostic value of these diagnostic tests, which will inform the development of recommendations regarding relevant tests to perform during a clinical assessment, and regarding the value of ultrasound scan findings in making decisions regarding treatment and referral. The prognostic model will be based on self-report information only, which will allow the future screening tool to be used during remote as well as face-to-face consultations with healthcare professionals.

### Ethics and Dissemination

The PANDA-S study has ethical approval from Yorkshire and The Humber – Sheffield Research Ethics Committee (18/YH/0346, IRAS Number: 242750). Results will be disseminated through peer-reviewed publications, social and mainstream media, professional conferences, and the patient and public involvement and engagement group supporting this study and through newsletters, leaflets and posters in participating sites.

## Supporting information

Figure 1 Flowchart

## Data Availability

Any requests for access to the data from anyone outside of the research team (e.g. collaboration, joint publication, and data sharing requests from publishers) will follow the Keele CTUs data sharing procedure.

## Acknowledgements

The authors acknowledge the support of the NIHR Clinical Research Networks: West Midlands, Thames Valley and South Midlands, and North West Coast and Keele Clinical Trials Unit for their support in delivering and hosting this research.

## Author contributions

Design: DAvdW, GW-J, CL, AH, HM, SH, BS, MB, SJ, RR, SW, CH, JC, TP, CM, ER, NEF, DB,JL, AHi

Data collection and project administration: DAvdW, GW-J, AH, HM, SH, SW, JC Writing original draft: GW-J, DAvdW, HM, BS, MB, SJ

Writing-review and editing, and review of final draft: DAvdW, GW-J, CL, AH, HM, SH, BS, MB, SJ, RR, SW, CH, JC, TP, CM, ER, NEF, DB,JL, AHi

