## Supplementary material for "Predicting pain and function outcomes in people consulting with shoulder pain: The PANDA-S clinical cohort and qualitative study protocol (ISRCTN 46948079)": Figure 1 Flowchart

**Figure 1: Study summary flow chart**

Approximately 40 [EMIS / SystmOne] General Practices in CRN West Midlands, North West Coast, and Thames Valley & South Midlands, alongside self-referral physiotherapy services [EMIS community] and Physiotherapy waiting lists

3. Physiotherapy waiting list

2. Self-referral Physiotherapy clinics (EMIS community web)

1. GP consultation Pop-up (EMIS / SystmOne)

**EITHER:** GP completes pop-up during consultation: indicates that patient consents to Keele CTU having their contact details

**OR:** GP completes pop-up after the consultation: weekly search to identify eligible patients

Auto-populated contact form e-mailed to Keele CTU (nhs e-mail)

Pop-up information downloaded and sent to Keele CTU

Referrals triaged against PANDA-S eligibility criteria

Physiotherapist confirms that patient consents to Keele CTU having their contact details (Auto-population of template embedded in EMIS community)

For those deemed eligible:

Study pack sent by Physio service*

Study pack* sent by Keele CTU; GP or Physio service

Consent, eligibility and questionnaire returned to Keele CTU

**EITHER:** Text sent by GP inviting patient to contact Keele CTU for further details

**OR:** Study pack* sent by GP

Additional consent given for medical record extraction and review

[**Optional**] Medical record extraction and review

Letter of invitation and separate Participant Information Leaflets to research clinic and to complete short-term follow-up (app / SMS)

Participant to telephone Keele CTU for research clinic appointment. If no response within 2 weeks: telephone call from Keele CTU to participant to check whether a clinic appointment is required

Participant to complete and return reply slip to take part in short-term data collection

Letter of invitation and Participant Information Leaflet (to purposive sample) for qualitative interview. Participant to complete and return reply slip to take part in interview

GPs from participating practices invited to qualitative interview (if patient-participant consents to this during their interview)

[**Optional**] Pain app / text every week for 3 months

Follow-up postal questionnaires: 3m, 6m, 12m, 24m, 36m (for detail of process, refer to study follow-up flow charts in the Appendix 14.1)

[**Optional**] Research Clinic: Consent; Clinical interview and physical assessment of the shoulder; Ultrasound scan of the shoulder

[**Optional**] Qualitative interview

Yes

Yes

Yes

No

No

No

*Letter of invitation; Participant Information Leaflet; Consent form; Eligibility screen; Baseline questionnaire; Return envelope
